# Inequalities in access to and outcomes of cardiac surgery in England: retrospective analysis of Hospital Episode Statistics (2010-2019)

**DOI:** 10.1101/2024.05.10.24307170

**Authors:** Florence Y Lai, Ben Gibbison, Alicia O’Cathain, Enoch Akowuah, John Cleland, Gianni D Angelini, Christina King, Gavin J Murphy, Maria Pufulete

**Author notes:** Correspondence to: Dr Maria Pufulete, Bristol Heart Institute, University of Bristol. Abbreviations*: CABG, Coronary artery bypass grafting; HES, Hospital Episode Statistics; NHS, National Health Service; ONS, Office for National Statistics; UK, United Kingdom.

## Abstract

**Objectives:** To characterise the variation in access to and outcomes of cardiac surgery for people in England.

**Methods:** We included people >18 years of age with a hospital admission for ischaemic heart disease (IHD) and heart valve disease (HVD) between 2010 and 2019. Within these populations, we identified patients who had coronary artery bypass graft (CABG) and/or valve surgery, respectively. We fitted logistic regression models to examine the effects of age, sex, ethnicity and socio-economic deprivation on having access to surgery and in-hospital mortality, 1-year mortality and hospital readmission.

**Results:** We included 292,140 people, of whom 28% were women, 11% were from an ethnic minority and 17% were from the most deprived areas. Across all types of surgery, 1 in 5 patients are readmitted to hospital within 1 year, rising to almost 1 in 4 for valve surgery patients. Women, Black people and people living in the most deprived areas were less likely to have access to surgery (CABG: 59%, 32%, 35% less likely; valve: 31%, 33%, 39% less likely, respectively) and more likely to die within 1 year of surgery (CABG: 24%, 85%, 18% more likely, respectively; valve: 19% (women) and 10% (people from most deprived areas) more likely).

**Conclusion:** Female sex, Black ethnicity, and economic deprivation are independently associated with limited access to cardiac surgery and higher post-surgery mortality. Actions are required to address these inequalities.

**WHAT IS ALREADY KNOWN ON THIS TOPIC:** There has been a marked improvement in short term (in hospital) outcomes of people having cardiac surgery. It is not clear if this translates to improvement in mid- and long-term outcomes and how these differ by demographic and socio-economic factors.

**WHAT THIS STUDY ADDS:** Across all types of cardiac surgery, 1 in 5 patients are readmitted to hospital within 1 year of surgery, rising to almost 1 in 4 for valve surgery patients.

While in hospital mortality has decreased markedly since 2010 (by 20%), 1 year mortality and hospital readmission remained largely unchanged.

Women, people of Black ethnicity and people from the most socially deprived areas were less likely to be offered cardiac surgery and more likely to die and be readmitted to hospital in the year after surgery.

**HOW THIS STUDY MIGHT AFFECT RESEARCH, PRACTICE OR POLICY:** Targeted interventions are required across the cardiovascular medicine pathway to improve mid- and long-term outcome in patients having cardiac surgery, addressing inequalities.

## Introduction

Cardiac surgery is high volume surgery, with about 28,000 adults a year undergoing a cardiac surgical procedure in the UK.^1^ It is the one of the costliest interventions carried out to treat cardiovascular disease by the UK National Health Service (NHS).^2^ While the National Adult Cardiac Surgery Audit (NACSA) has shown a steady improvement in short term outcomes of people having cardiac surgery, mid- and long-term outcomes are unknown.

The influence of social determinants of health (e.g., gender, ethnicity and deprivation) on mid and long-outcomes of cardiac surgery have not been explored. It is known that women and people from ethnic minorities and of low socio-economic status have worse short term outcomes after all types of cardiac surgery (in-hospital), ^3–7^ but it is unclear whether these characteristics also translate to poorer mid (one year) and long-term outcomes (three and five years respectively).

Our study, using the Hospital Episode Statistics (HES) and UK Office for National Statistics data, has two objectives: to describe mid- and long-term outcomes after cardiac surgery, previously not described in the UK, and characterise the variation in utilisation and outcome of cardiac surgery by sex, ethnicity and deprivation, whilst also examining intersections between these characteristics.

## Methods

### Study design

Retrospective cohort study using Hospital Episode Statistics (HES, England) linked with Office for National Statistics (ONS) mortality data

### Data sources

HES covers all admissions to NHS hospitals or independent providers that are funded by NHS.^8^ Each anonymised record contained demographics (age, sex, ethnicity, area of residence, index of multiple deprivation, IMD), administrative (admission and discharge dates, admission method, discharge destination, etc.) and clinical information (diagnoses and procedures performed). Diagnoses were coded based on the International Classification of Diseases, Tenth Revision (ICD-10), and procedures were coded by Office of Population Censuses and Surveys Classification of Interventions and Procedures (OPCS-4). Our dataset comprised all hospital admissions for two cardiovascular diseases: ischaemic heart disease (IHD) and heart valve diseases (HVD) which require cardiac surgery as treatment in England between April 2010 and March 2019 (financial years in the NHS in England).

### Study populations

All adult patients (>18 years) who had at least one hospital admission (not including day cases) with a IHD diagnosis and a HVD diagnosis respectively in each financial year during 2010 to 2019. Within these populations, we further identified patients who had cardiac surgery defined as coronary artery bypass graft (CABG) and/or valve surgery. When examining the outcomes of cardiac surgery patients, we included each patient’s first episode of cardiac surgery in the study period and excluded those who had undergone cardiac surgery in the previous two years. The ICD-10 and OPCS-4 codes used to define the study populations as well as CABG and valve surgery are shown in *Supplementary Table S1*.

Ethnicity was grouped as White (British, Irish and other White background), Black (Caribbean, African and other Black background), South Asian (Bangladeshi, Indian, Pakistani), and others (mixed and other ethnic background). Socio-economic status was defined using area-level Index of Multiple Deprivation (IMD) 2015 ^9^ scores. IMD quintiles were created by dividing the deprivation scores of individual areas into fifths ranging from the 20% most deprived areas to the 20% least deprived areas.

In order to adjust for pre-existing poor health, we identified comorbidities (Royal College of Surgeons (RCS) Charlson Score ^10^ based on diagnoses recorded within 1 year prior to the index admission) and frailty status ^9^ (defined as occurrence of one or more of seven domains associated with frailty using diagnoses recorded in all hospital admissions within 2 years before the index admission) (*Supplementary Tables S2 and S3*).

### Study outcomes

For the IHD and HVD populations, we calculated the utilisation of cardiac surgery, defined as the proportion of IHD patients who underwent CABG surgery and the proportion of HVD patients who underwent heart valve surgery.

For the cardiac surgery population, we calculated mortality (in-hospital and at 1, 3 and 5 years) and hospital readmission for: cardiovascular causes, heart failure and stroke / transient ischaemic attack. Readmission for cardiovascular causes was defined as admission to any NHS hospitals post cardiac surgery with a cardiovascular disease (ICD10 I00-I99) as the primary diagnosis. Readmission for heart failure and stroke/TIA was defined using both primary and secondary diagnoses.

### Statistical analysis

To describe access to surgery over time in the IHD and HVD populations, we calculated rates of CABG surgery per 1000 IHD patients and rates of valve surgery per 1000 HVD patients, standardised by age, Charlson index and frailty status using the respective patient populations in 2019 as the standard population. We examined standardised rates separately for the following subgroups: CABG surgery and valve surgery, by sex, ethnic group and IMD quintile. We fitted two logistic regression models to examine the effects of age, sex, ethnicity and IMD (adjusted for covariates including Charlson comorbidities, frailty status, year of surgery) on having access to CABG in the IHD population and having access to valve surgery in the HVD population.

To assess outcomes from surgery, we separated the patient cohorts into 2010-2014 and 2015-2019. We used the earlier cohort for calculating 1, 3, and 5-year outcomes and the later cohort for calculating 1-year outcomes. We fitted logistic regression models for in-hospital mortality and 1-year outcomes only to examine the effects of sex, ethnicity and IMD adjusted for age, Charlson index, frailty status, year of surgery, and operative characteristics (elective/emergency surgery, and whether the surgery involved cardiopulmonary bypass). Models were fitted separately for cardiac surgery patients undergoing CABG alone, valve surgery alone and combined CABG and valve surgery.

## Results

The characteristics of the 2010-2014 and 2015-2019 populations of cardiac surgery patients are shown in **Table 1**. In 2010-2014, there were 143,104 patients (54% isolated CABG, 32% isolated valve surgery and 14% combined CABG and valve). In 2015-2019, there were 149,036 patients (45% isolated CABG, 43% isolated valve surgery and 12% combined CABG and valve). Standardised rates of both CABG and valve surgery decreased between 2010-2019 (*Supplementary Figure S1*), from 30.1 to 24.8 per 1000 IHD patients and from 87.9 to 69.9 per 1000 HVD patients, respectively. The demographics of the two populations (age, sex, ethnicity, socio-economic quintile) were similar, although the proportions of multi-morbid and frail patients and emergency admissions were higher in 2015-2019, across all types of surgery reflecting the increasing comorbidity and frailty over time in the admitted IHD and HVD populations (*Supplementary Tables S4 and S5*).

**Table 1.**
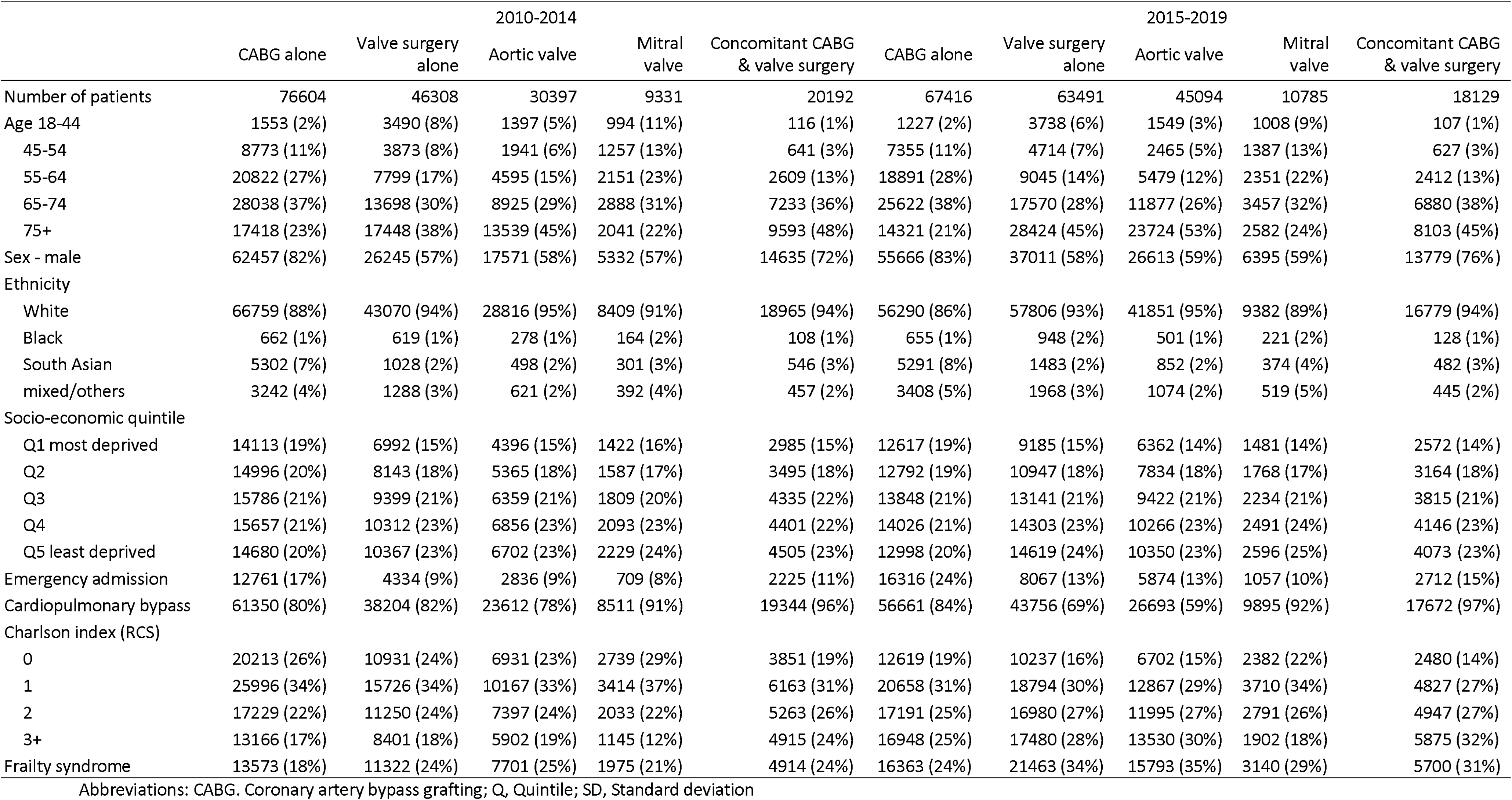
Characteristics of cardiac surgery patients across two time periods (2010-2014 and 2015-2019)

### Access to cardiac surgery

The adjusted associations between age, sex, ethnicity and socio-economic quintile on access to CABG and valve surgery are shown in **Figure 1**. Women were less likely to have CABG and valve surgery than men (OR 0.41, 95% CI 0.41-0.42 and OR 0.69, 95% CI 0.69-0.70). Compared with White people, Black people were less likely to have surgery (CABG: OR 0.68, 95% CI 0.64-0.71; Valve: OR 0.67, 95% CI0.64-0.70), while South Asian people were more likely to have CABG (OR 1.49, 95% CI1.46-1.52) but not valve surgery (OR 0.72, 95% CI 0.69-0.75). For both CABG and valve surgery, there was almost a linear association between increasing level of deprivation and decreasing odds of getting surgery, with people in the most deprived group being 35% and 39% less likely to have CABG and valve surgery, respectively, than people in the least deprived group (OR 0.65, 95% CI 0.64-0.68 and OR 0.61, 95% CI 0.60-0.62). For intersectionality, standardised rates of both CABG and valve surgery were lowest in Black women and women from the lowest socio-economic quintile, with little change to this pattern between 2010 and 2019 (*Supplementary Figure S2*).

**Figure 1.**
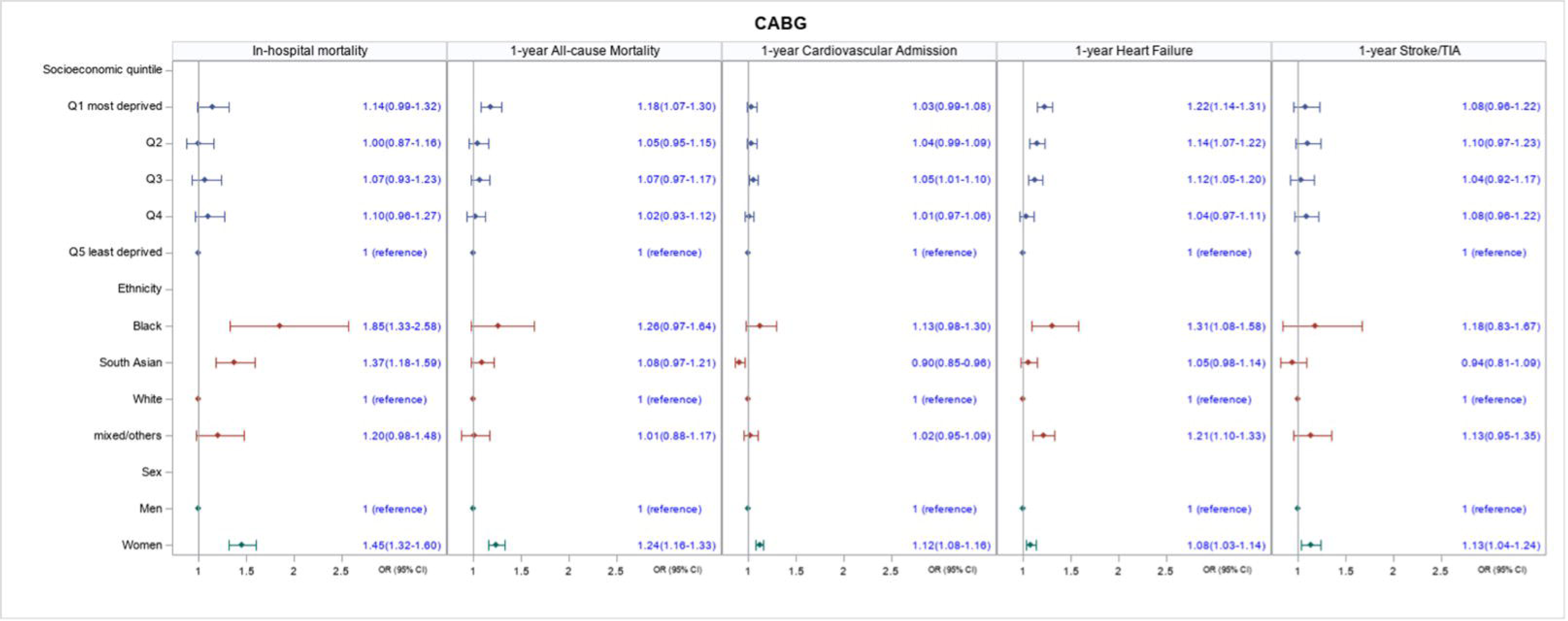
Association of age, sex, ethnicity and socio-economic quintile on having access to (left) CABG in patients with IHD and (right) valve surgery in patients with heart valve disease All models are adjusted for age, Charlson index, frailty status, year of surgery, and operative characteristics (elective/emergency surgery, and whether the surgery involved cardiopulmonary bypass). CABG: coronary artery bypass grafting; IHD: ischaemic heart disease, HVD: heart valve disease.

### Outcomes from Cardiac Surgery

#### In-hospital mortality (short term)

In hospital mortality decreased across all types of cardiac surgery between 2010-2014 and 2015-2019 and was lowest in CABG (1.7% and 1.4%, respectively) and highest in combined CABG and valve surgery (5.4% and 4.5% respectively, **Table 2**).

**Table 2.** Outcomes of patients undergoing CABG and valve surgery in 2010-2019.

|  | 2010-2014 |  |  |  |  | 2015-2019 |  |  |  |  |
| --- | --- | --- | --- | --- | --- | --- | --- | --- | --- | --- |
|  | CABG<br>alone | Valve<br>surgery<br>alone | Aortic<br>valve | Mitral<br>valve | Concomitant<br>CABG & valve<br>surgery | CABG<br>alone | Valve<br>surgery<br>alone | Aortic<br>valve | Mitral<br>valve | Concomitant<br>CABG & valve<br>surgery |
| No of patients | 76604 | 46308 | 30397 | 9331 | 20192 | 67416 | 63491 | 45094 | 10785 | 18129 |
| In-hospital mortality | 1.7% | 3.0% | 2.5% | 2.6% | 5.4% | 1.4% | 2.3% | 1.8% | 2.0% | 4.5% |
| Mortality (all-cause) |  |  |  |  |  |  |  |  |  |  |
| 1 year | 3.8% | 7.6% | 7.5% | 5.4% | 10.5% | 3.4% | 7.5% | 7.8% | 4.9% | 9.1% |
| 3 year | 7.6% | 14.7% | 15.8% | 9.4% | 17.8% |  |  |  |  |  |
| 5 year | 12.6% | 22.7% | 25.2% | 13.9% | 26.8% |  |  |  |  |  |
| Cardiovascular-cause hospital readmission |  |  |  |  |  |  |  |  |  |  |
| 1 year | 17.3% | 25.4% | 23.6% | 27.0% | 26.6% | 15.5% | 23.4% | 22.1% | 25.1% | 24.0% |
| 3 year | 27.2% | 34.5% | 32.8% | 35.6% | 36.3% |  |  |  |  |  |
| 5 year | 34.7% | 41.3% | 39.9% | 41.6% | 43.7% |  |  |  |  |  |
| Hospital admission for heart failure |  |  |  |  |  |  |  |  |  |  |
| 1 year | 6.4% | 11.7% | 11.2% | 10.4% | 13.0% | 8.0% | 14.9% | 14.7% | 13.5% | 14.8% |
| 3 year | 10.5% | 18.4% | 18.1% | 16.2% | 19.9% |  |  |  |  |  |
| 5 year | 15.0% | 24.3% | 24.2% | 21.0% | 26.5% |  |  |  |  |  |
| Hospital admission for stroke/TIA |  |  |  |  |  |  |  |  |  |  |
| 1 year | 2.1% | 3.4% | 3.5% | 3.0% | 4.0% | 2.0% | 3.8% | 4.1% | 2.9% | 4.2% |
| 3 year | 4.0% | 5.9% | 6.2% | 5.3% | 6.8% |  |  |  |  |  |
| 5 year | 5.9% | 8.1% | 8.5% | 7.0% | 9.3% |  |  |  |  |  |

Abbreviations: CABG, Coronary artery bypass grafting.

Across all types of surgery, women had a higher in-hospital mortality rate than men (CABG: OR 1.45, 95% CI 1.32-1.60; valve surgery: OR 1.31, 95% CI 1.21-1.4; CABG and valve surgery: OR 1.50, 95% CI 1.36-1.66, **Table 2** and **Figures 2-4**). In terms of ethnicity, South Asian patients had a higher in-hospital mortality compared with White patients (CABG: OR 1.37, 95% CI 1.18-1.59, valve surgery: OR 1.53, 95% CI 1.23-1.89; CABG and valve surgery: OR 1.67, 95% CI 1.33-2.10). Black patients had higher in-hospital mortality than White patients after CABG (OR 1.85, 95% CI 1.33-2.58), but not valve or CABG and valve surgery. Although crude in-hospital mortality was higher in patients with the highest socio-economic deprivation vs. those with the lowest socio-economic deprivation across all types of surgery (**Table 2**), these differences were not significant after adjustment (**Figures 2-4).**

**Figure 2.**
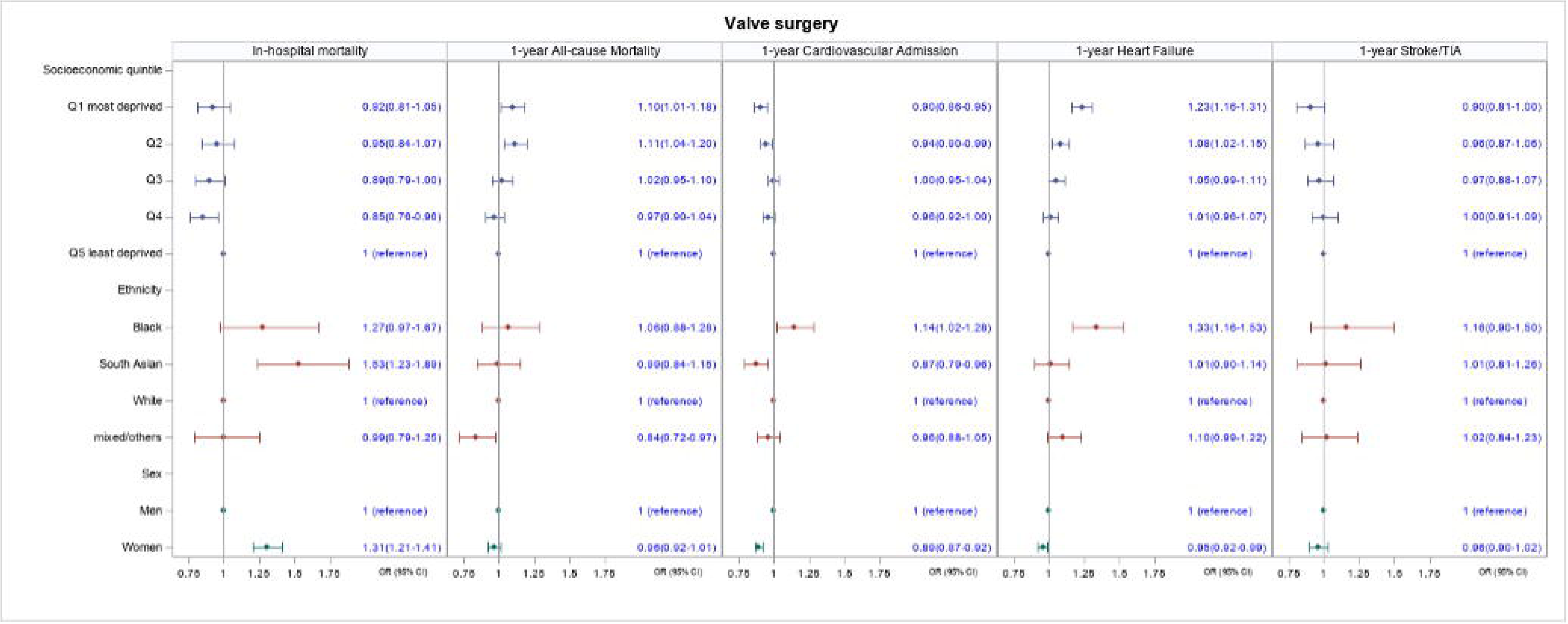
Association of sex, ethnicity and socio-economic quintile with in-hospital mortality and 1-year outcomes for patients undergoing coronary artery bypass grafting (CABG) All models are adjusted for age, Charlson index, frailty status, year of surgery, and operative characteristics (elective/emergency surgery, and whether the surgery involved cardiopulmonary bypass)

#### Mid- and long-term outcomes

At 1 year, mortality and hospital readmissions decreased slightly (by 8% and 9%, respectively) across the two time periods (2010-2014 and 2015-2019) (**Table 2**). Mortality was higher in patients undergoing valve surgery than CABG (7.6% valve surgery vs 3.8% CABG in 2010-2014, 7.5% vs 3.4% in 2015-2019). By 5 years, mortality had generally trebled across all types of surgery (to between 13% and 27%, 2010-2014).

At 1 year, readmission for any cardiovascular cause was higher in patients undergoing valve surgery than CABG (25.4% vs 17.3% in 2010-2014 and 23.4% vs 15.5% in 2015-2019). By 5 years, readmission rates had more than doubled across all types of surgery, with between 35% and 44% of all cardiac surgery patients having a readmission (2010-2014).

Crude mortality and hospital readmission rates at 1 year were generally higher in women compared with men across all types of surgery (*Supplementary Table S6*). Mortality differences persisted for all outcomes after adjustment in women who had CABG (**Figure 2**) and combined CABG and valve (**Figure 4**) who had a 24% and 19% increased odds of death (OR 1.24, 95% CI 1.16-1.33 and OR 1.19, 95% CI 1.11-1.29), respectively, compared with men. Women who had CABG surgery had increased odds of hospital readmission (OR 1.12, 95% CI 1.08-1.16) compared with men, but this was not the case for women who had valve surgery (**Figure 3** and **Figure 4**).

**Figure 3.**
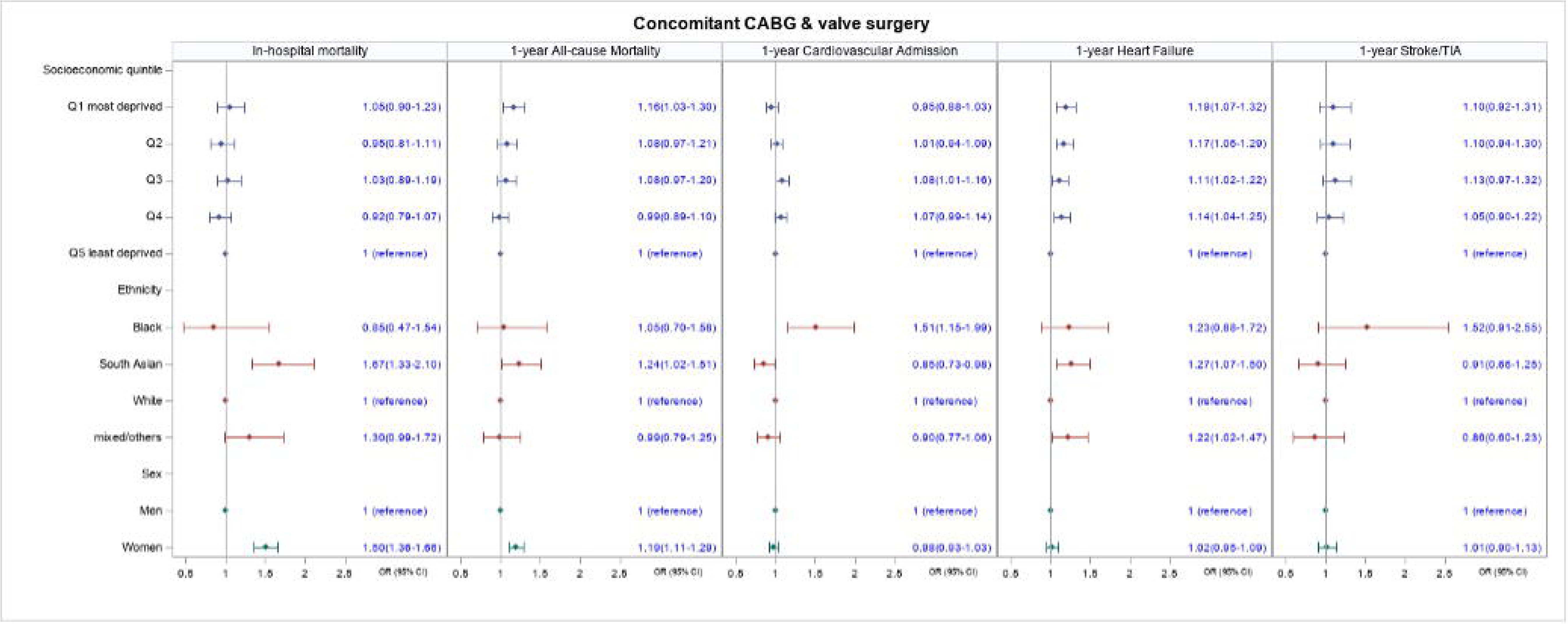
Association of sex, ethnicity and socio-economic quintile with in-hospital mortality and 1-year outcomes for patients undergoing alone valve surgery All models are adjusted for age, Charlson index, frailty status, year of surgery, and operative characteristics (elective/emergency surgery, and whether the surgery involved cardiopulmonary bypass)

Crude 1 year mortality and hospital readmission were generally higher in Black patients compared with White patients across all types of cardiac surgery (*Supplementary Table S6*). After adjustment, there were no significant differences in 1 year mortality between Black and White patients, although Black CABG and valve surgery patients had a 31% and 33%, respectively, higher odds of being readmitted to hospital for heart failure (OR 1.31, 95% CI 1.08-1.58 and OR 1.33, 95% CI 1.16-1.53, respectively); and Black valve and combined CABG and valve patients had a 14% and 51% higher odds, respectively, of being readmitted to hospital for any cardiovascular cause (OR 1.14, 95% CI 1.02-1.28 and OR 1.51, 95% CI 1.15-1.99, respectively).

South Asian patients undergoing isolated CABG and valve surgery had similar 1 year mortality as White patients (**Figure 2** and **Figure 3**), but those undergoing CABG and valve surgery had higher odds of dying at 1 year (OR 1.24, 95% CI 1.02-1.51). Compared with White patients, South Asian patients had a slightly lower odds of readmission for any cardiovascular cause (CABG: OR 0.90, 95% CI 0.85-0.96, valve surgery: OR 0.87, 95% CI 0.79-0.96; combined CABG and valve surgery: OR 0.85 95%CI 0.73-0.98) but higher odds of readmission for heart failure (OR 1.27, 95% CI 1.07-1.50).

The odds of dying increased with increasing level of deprivation (**Figures 2-4**). Across all types of surgery, patients in the most deprived quintiles had a 18% (CABG), 10% (valve) and 16% (combined CABG and valve) increased odds of dying by 1 year compared with patients in the least deprived quintile (OR 1.18, 95% CI 1.07-1.30; OR 1.10, 95% CI 1.01-1.18; OR 1.16, 95% CI 1.03-1.30, respectively). Similarly, the odds of heart failure readmission increased with increasing level of deprivation: CABG by 22% (OR 1.22, 95% CI 1.14-1.31); valve surgery by 23% (OR.1.23, 95% CI 1.16-1.31); combined CABG and valve surgery by 19% (OR 1.19, 95% CI 1.07-1.32).

**Figure 4.**
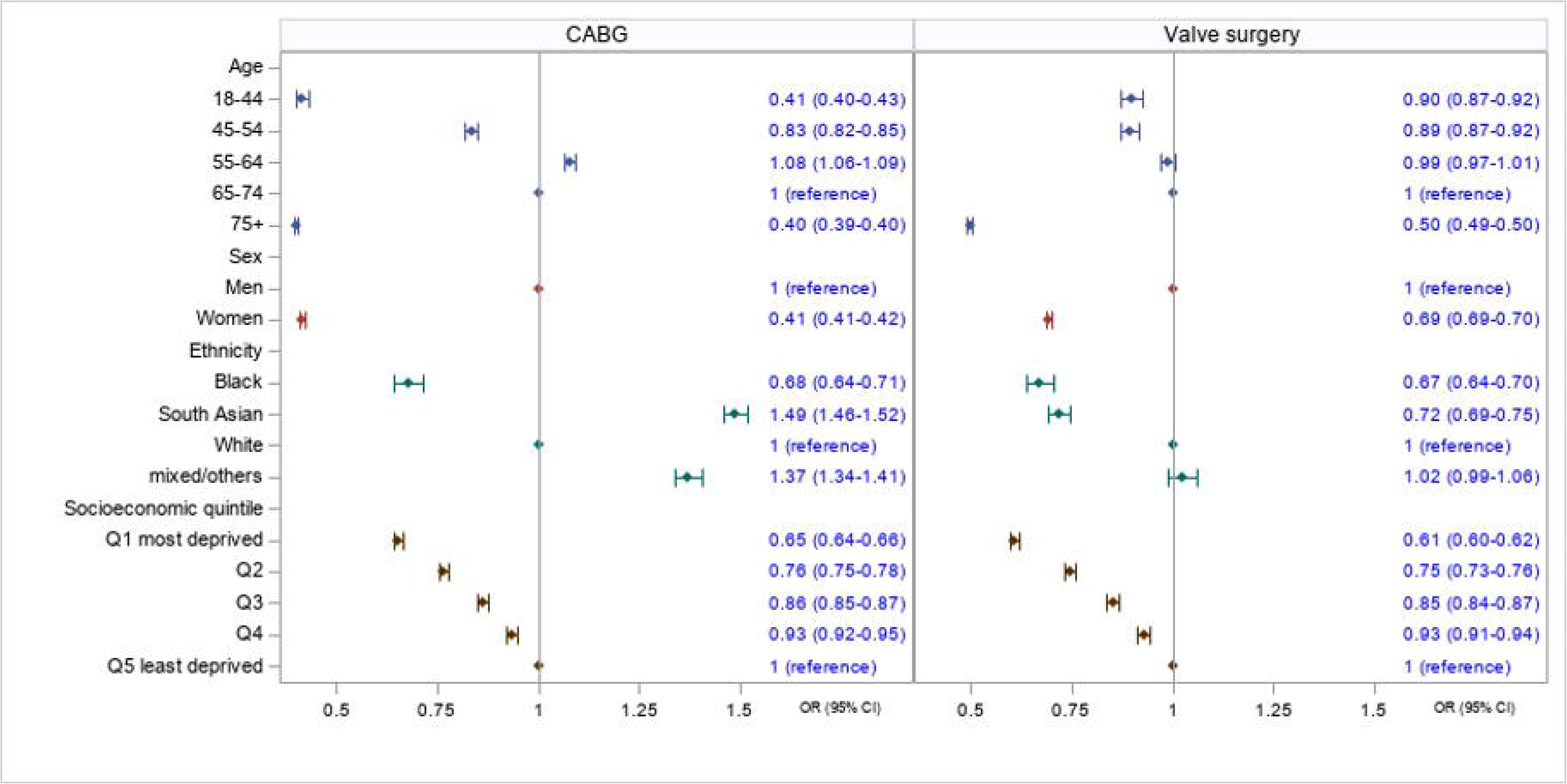
Association of sex, ethnicity and socio-economic quintile with in-hospital mortality and 1-year outcomes for patients undergoing concomitant coronary artery bypass grafting (CABG) and valve surgery All models are adjusted for age, Charlson index, frailty status, year of surgery, and operative characteristics (elective/emergency surgery, and whether the surgery involved cardiopulmonary bypass)

## Discussion

There has been a decline in the use of cardiac surgery as a treatment modality over time, a pattern observed in both Europe^1^ and the US^11^. The increased risk profile of patients undergoing cardiac surgery with the concurrent decrease in in-hospital mortality^12^ reflects the improved safety and quality of cardiac surgery. However, mid-term (1 year) outcomes (mortality and readmission) have not seen the same proportional improvement. Across all types of cardiac surgery, 1 in 5 patients are readmitted to hospital within 1 year of surgery, rising to almost 1 in 4 for valve surgery patients.

This suggests that the biggest improvements to care have been in the in-hospital surgical pathways rather than “joined-up” care. Cardiac surgery is often used as an example of an efficient patient centred care pathway that achieves better outcomes than many other types of major surgery,^13^ yet it is clear from these data that much remains to be done in implementing a complete model of care that optimises both the pre-operative selection and optimisation of patients and the postoperative period after hospital discharge.

We observed disparities in access to surgery and outcomes of surgery by demographic and socio-economic characteristics, with women, people from deprived backgrounds and people of Black ethnicity being less likely to be offered surgery and experiencing poorer outcomes. For deprivation, there was an almost linear pattern of decreasing odds of getting surgery with increasing level of deprivation, even after adjustment for multiple covariates, including frailty and comorbidities. From an intersectional perspective, the disparities widened when more than one inequality factor was considered; for example, women from deprived areas were less likely to get surgery than men from deprived areas, while Black women were less likely to get surgery than Black men.

The reasons for the differences in outcomes between women and men after cardiac surgery, in particular CABG, have been extensively discussed.^13^ Compared with men, women have delayed diagnosis, more comorbidities, a broader array of symptoms,^14^ more unstable or acute presentation, a higher rate of small vessel disease and reduced patient and clinician perception of actual risk.^13,15,16^ Some of these factors also likely apply to Black people and people from deprived backgrounds.^17,18^

Ethnic inequalities in the NHS have been attributed to poor education, social status, and poverty outside the health system. Inside the health system, there may be poor quality or discriminatory treatment from healthcare staff, a lack of high quality recorded ethnic monitoring data; lack of appropriate interpreting services^19^ and avoidance of seeking help for health problems due to fear of racist treatment from healthcare professionals. Whilst in the US, one of the key mechanisms of ethnic inequalities in cardiac surgery has been purported to be lack of access to high-quality hospitals for Black ethnic groups^20^, this is not true in the UK. Cardiac surgery is a specially commissioned, nationally funded service and the vast majority of cardiac surgery takes place in these regional centres. Whilst clustering of ethnic groups within particular centres may occur, these are often high-volume, high-quality centres – the causes are clearly wider than poor quality care alone. There is an urgent need to address inequalities through enhanced data linkage and improved transparency and publication of data from benchmarking exercises by inequality characteristics and ensuring equity of workforce and pathways patients use to access care.

## Limitations

A key limitation of using HES is that it relies on diagnostic and procedure codes for identifying patients and outcomes. Coding practices vary between hospitals, particularly for diagnostic coding and although the accuracy of coding has improved markedly over the past 20 years^21^, and caution is needed when interpreting time-series of HES data. However, our findings, in terms of trends in cardiac surgery, increased complexity of patients and short-term (in hospital) outcomes mirror finding from the National Adult Cardiac Surgery Audit,^1^ which uses data input directly by clinicians, therefore our data are likely to be broadly accurate.

Another limitation is that over 10% of patients within the HES database have an unknown ethnicity^22^, therefore our findings with regards with ethnicity need to be interpreted with caution. We did not attempt to impute missing ethnicity data, largely because the value of this analysis is uncertain given that ethnicity data are not likely to be missing at random and there are likely to be fundamental differences between the population with and without ethnicity status. Also, the “mixed/other” ethnic group is heterogeneous and cannot be meaningfully interpreted because it includes people of mixed race and specific ethnic groups (e.g., Chinese) for which numbers were too small to analyse separately.

We used patients admitted to hospital with ischaemic heart disease and valve disease as our denominators for CABG and valve surgery rates. These populations do not necessarily represent the population “at-risk” of having cardiac surgery – there are likely to be underlying differences in eligibility for surgery based not only on demographics and comorbidities (for which we adjusted) but also pathology (e.g., greater proportion of degenerative valve disease in the admitted population vs. the valve surgery population) and other elements of operative risk scores (e.g., EuroSCORE II^23^ / STS risk-score ^24^) which are not collected in HES.

## Conclusions

The improvements in in-hospital outcomes after cardiac surgery that have been so well documented over the past decade do not translate into a similar improvement in mid- and long-term outcomes. Whilst for some groups of cardiac surgery patients, both absolute and relative inequalities have decreased, this pattern is not consistent. These are stark in the case of Black people, South Asians and women who are consistently more likely to die after heart surgery than their white and male peers. Targeted interventions are required across the cardiovascular medicine pathway to ensure that interventions are applicable to and implemented in underserved populations. Healthcare providers must also undertake surveillance using routinely collected data to ensure that their interventions are reaching underserved groups ^25^.

## Supporting information

Supplementary Analysis

## Data availability statement

The data are not available for sharing because the data were acquired under a data sharing agreement from NHS Digital, for which conditions of use (and further use) apply.

## Patient consent for publication

Not applicable.

## Ethics approval

This study does not need consent from individuals included in the HES dataset because the data are pseudonymised (to the level required by ISB1523 Anonymisation Standard for Publishing Health and Social Care Data) for analysis by NHS Digital and the analysis and presentation of data follows current NHS Digital guidance for the use of HES data for research purposes.

## Contributors

FYL designed the study and undertook the statistical analysis. BG interpreted the data and co-wrote the manuscript. AOC contributed to the discussion on inequalities and revised the manuscript for intellectual content. EA, GDA and GM provided cardiac surgery expertise and revised the manuscript for intellectual content. JC provided cariology expertise and revised the manuscript for intellectual content. CK provided the PPI perspective. MP designed and obtained funding for the study, interpreted the data and co-wrote the manuscript.

## Disclaimer

The funder had no role in the conception, design or conduct of the study; collection, analysis or interpretation of the data; writing of the manuscript; or decision to submit the manuscript for publication.

## Patient and public involvement

A PPI group of cardiac surgery patients was assembled specifically for the NIHR Programme Development Grant (NIHR203304). The group was diverse, including individuals from all types of cardiac surgery and most demographic groups investigated in this research. PPI members met regularly to discuss and provide input into the study and its interpretation from the patient perspective and the PPI lead (Christina King) is an author on this manuscript.

