## Supplementary Analysis for "Inequalities in access to and outcomes of cardiac surgery in England: retrospective analysis of Hospital Episode Statistics (2010-2019)"

**Supplementary material**

| **Supplementary Table/Figure** | | Page |
| --- | --- | --- |
| *Supplementary Table S1* | ICD-10 and OPCS-4 codes used to define the study populations, patients' comorbidities (RCS Charlson Score) and frailty status | 2 |
| *Supplementary Table S2* | Comorbidities (Charlson index RCS) and frailty of admitted ischaemic heart disease and those undergoing CABG surgery in England, 2010-2019 | 3 |
| *Supplementary Table S3* | Comorbidities (Charlson index RCS) and frailty of admitted heart valve diseases patients and those undergoing heart valve surgery in England, 2010-2019 | 4 |
| *Supplementary Figure S1* | Number and Standardised Rates of CABG and valve surgery in England, UK 2010-2019 | 5 |
| *Supplementary Figure S2* | Standardised rates of CABG and valve surgery per 1000 admitted patients with ischaemic heart disease (IHD) and heart valve disease (HVD) | 6 |
| *Supplementary Table S4* | Characteristics of admitted patients with ischaemic heart disease (IHD) and those undergoing CABG in England, 2010-2019 | 7 |
| *Supplementary Table S5* | Characteristics of admitted patients with heart valve diseases (HVD) and those undergoing valve surgery in England, 2010-2019 | 8 |
| *Supplementary Table S6* | Outcomes of patients undergoing CABG and valve surgery in 2010-2019 by sex, ethnic group and socio-economic quintile | 9 |

| **Supplementary Table S1. ICD-10 and OPCS-4 codes used to define the study populations, patients' comorbidities (RCS Charlson Score) and frailty status** | | |
| --- | --- | --- |
| **Population** | **Coding** |  |
| Ischaemic heart disease | ICD10 | I20-I25 |
| Heart valve disease | ICD10 | I01, I05-I08, I33-I39, I511, I512 |
| CABG^*^ | OPCS-4 | K40-K46 |
| Heart valve surgery^*^ | OPCS-4 | K25-K31, K34, K36.1, K36.2 |
| Aortic valve surgery | OPCS-4 | K26, K30.2, K31.2 |
| Mitral valve surgery | OPCS-4 | K25, K30.1, K31.1, K34.1 |
| *Charlson index (RCS)* | | |
| Malignancy | ICD10 | C00–C26, C30–C34, C37–C41, C43, C45–C58, C60–C76, C80–C85, C88, C90–C97 |
| Metastatic solid tumour | ICD10 | C77–C79 |
| Myocardial infarction | ICD10 | I21, I22, I23, I252 |
| Congestive cardiac failure | ICD10 | I11, I13, I255, I42, I43, I50, I517 |
| Peripheral vascular disease | ICD10 | I70–I73, I770, I771, K551, K558, K559, R02, Z958, Z959 |
| Cerebrovascular disease | ICD10 | G45, G46, I60–I69 |
| Dementia | ICD10 | A810, F00–F03, F051, G30, G31 |
| Chronic pulmonary disease | ICD10 | I26, I27, J40–J45, J46, J47, J60–J67, J684, J701, J703 |
| Rheumatological disease | ICD10 | M05, M06, M09, M120, M315, M32–M36 |
| Liver disease | ICD10 | B18, I85, I864, I982, K70, K71, K721, K729, K76, R162, Z944 |
| Diabetes mellitus | ICD10 | E10–E14 |
| Hemiplegia or paraplegia | ICD10 | G114, G81–G83 |
| Renal disease | ICD10 | I12, I13, N01, N03, N05, N07, N08, N171, N172, N18, N19, N25, Z49, Z940, Z992 |
| AIDS/HIV infection | ICD10 | B20–B24 |
| *Frailty syndrome* | | |
| Dementia and delirium | ICD10 | F00-F05, G30, G311, G310, R41 |
| Mobility problems | ICD10 | R26, R298 |
| Fall and fractures | ICD10 | S32, S33, S42, S43, S52, S53, S62, S63, S72, S73, W00-W19, M80, M81, M966, R296, R54, R55 |
| Pressure ulcers and weight loss | ICD10 | L89, R634, R636, Z724 |
| Incontinence | ICD10 | R32, R15 |
| Dependence and care | ICD10 | Z74, Z75 |

AIDS: acquired immunodeficiency syndrome; CABG: coronary artery bypass grafting; HIV human immunodeficiency virus; ICD: International Classification of Diseases; OPCS: RCS: Royal College of Surgeons

^*^ Bortolussi G. McNulty D, Waheed H. et al. Identifying cardiac surgery operations in hospital episode statistics administrative database, with an OPCS-based classification of procedures, validated against clinical data. BMJ Open. 2019; 9(3): e023316. Published online 2019 Mar 23. doi: 10.1136/bmjopen-2018-023316

| **Supplementary Table S2**. Comorbidities (Charlson index RCS) and frailty of admitted ischaemic heart disease and those undergoing CABG surgery in England, 2010-2019 | | | | | | | | | | |
| --- | --- | --- | --- | --- | --- | --- | --- | --- | --- | --- |
|  | Admitted Ischaemic Heart Disease Patients | | | | | CABG patients | | | | |
| Year | 2010-2011 | 2012-2013 | 2014-2015 | 2016-2017 | 2018-2019 | 2010-2011 | 2012-2013 | 2014-2015 | 2016-2017 | 2018-2019 |
| N | 1190870 | 1206648 | 1224683 | 1234160 | 1285859 | 39511 | 38245 | 36774 | 34484 | 33031 |
| Charlson index (RCS) |  |  |  |  |  |  |  |  |  |  |
| 0 | 320424 (27%) | 259390 (21%) | 193816 (16%) | 177623 (14%) | 169583 (13%) | 10991 (28%) | 9102 (24%) | 7596 (21%) | 6247 (18%) | 5175 (16%) |
| 1 | 392838 (33%) | 372893 (31%) | 342830 (28%) | 326788 (26%) | 320892 (25%) | 13408 (34%) | 12596 (33%) | 11737 (32%) | 10431 (30%) | 9348 (28%) |
| 2 | 266377 (22%) | 294140 (24%) | 316602 (26%) | 317139 (26%) | 324616 (25%) | 8599 (22%) | 9137 (24%) | 9155 (25%) | 8843 (26%) | 8779 (27%) |
| 3+ | 211231 (18%) | 280225 (23%) | 371435 (30%) | 412610 (33%) | 470768 (37%) | 6513 (16%) | 7410 (19%) | 8286 (23%) | 8963 (26%) | 9729 (29%) |
| Malignancy | 102827 (9%) | 108002 (9%) | 114482 (9%) | 120147 (10%) | 129784 (10%) | 1232 (3.1%) | 1263 (3.3%) | 1296 (3.5%) | 1317 (3.8%) | 1258 (3.8%) |
| Metastatic solid tumour | 25011 (2%) | 27183 (2%) | 29233 (2%) | 31257 (3%) | 34737 (3%) | 68 (0.2%) | 82 (0.2%) | 80 (0.2%) | 86 (0.2%) | 94 (0.3%) |
| Myocardial infarction | 133636 (11%) | 293293 (24%) | 515176 (42%) | 525652 (43%) | 550501 (43%) | 9567 (24%) | 12475 (33%) | 14871 (40%) | 14055 (41%) | 13567 (41%) |
| Congestive cardiac failure | 230801 (19%) | 252819 (21%) | 279808 (23%) | 313414 (25%) | 357192 (28%) | 9852 (25%) | 9947 (26%) | 9884 (27%) | 10945 (32%) | 12138 (37%) |
| Peripheral vascular disease | 119175 (10%) | 131927 (11%) | 140611 (11%) | 149407 (12%) | 167241 (13%) | 6027 (15%) | 5875 (15%) | 5733 (16%) | 5967 (17%) | 6022 (18%) |
| Cerebrovascular disease | 110457 (9%) | 116887 (10%) | 123443 (10%) | 131855 (11%) | 149924 (12%) | 2757 (7.0%) | 2878 (7.5%) | 3167 (8.6%) | 3411 (9.9%) | 3879 (11.7%) |
| Dementia | 76053 (6%) | 90768 (8%) | 105841 (9%) | 115592 (9%) | 123901 (10%) | 137 (0.3%) | 192 (0.5%) | 171 (0.5%) | 191 (0.6%) | 198 (0.6%) |
| Chronic pulmonary disease | 299441 (25%) | 320549 (27%) | 344563 (28%) | 364397 (30%) | 398731 (31%) | 7075 (18%) | 7321 (19%) | 7021 (19%) | 6803 (20%) | 6736 (20%) |
| Rheumatological disease | 47252 (4%) | 51659 (4%) | 56843 (5%) | 62724 (5%) | 71899 (6%) | 1174 (3.0%) | 1197 (3.1%) | 1239 (3.4%) | 1240 (3.6%) | 1196 (3.6%) |
| Liver disease | 17927 (2%) | 20928 (2%) | 24990 (2%) | 31058 (3%) | 43999 (3%) | 373 (0.9%) | 474 (1.2%) | 502 (1.4%) | 616 (1.8%) | 904 (2.7%) |
| Diabetes mellitus | 310515 (26%) | 332966 (28%) | 359100 (29%) | 380410 (31%) | 413887 (32%) | 11142 (28%) | 11526 (30%) | 11684 (32%) | 11609 (34%) | 11707 (35%) |
| Hemiplegia or paraplegia | 21683 (2%) | 21522 (2%) | 22658 (2%) | 26370 (2%) | 29293 (2%) | 304 (0.8%) | 246 (0.6%) | 255 (0.7%) | 279 (0.8%) | 271 (0.8%) |
| Renal disease | 172413 (14%) | 194395 (16%) | 221136 (18%) | 243576 (20%) | 280205 (22%) | 3731 (9.4%) | 3534 (9.2%) | 3584 (9.7%) | 3764 (10.9%) | 4362 (13.2%) |
| Frailty syndrome | 427416 (36%) | 480077 (40%) | 532715 (43%) | 576561 (47%) | 642477 (50%) | 6747 (17%) | 7665 (20%) | 8099 (22%) | 8637 (25%) | 9373 (28%) |
| Mental | 122317 (10%) | 148342 (12%) | 174976 (14%) | 197329 (16%) | 218904 (17%) | 1217 (3.1%) | 1573 (4.1%) | 1684 (4.6%) | 1992 (5.8%) | 2096 (6.3%) |
| Mobility problems | 33433 (3%) | 65646 (5%) | 92850 (8%) | 116805 (9%) | 146991 (11%) | 338 (0.9%) | 532 (1.4%) | 591 (1.6%) | 795 (2.3%) | 984 (3.0%) |
| Falls and Fractures | 273356 (23%) | 296452 (25%) | 326891 (27%) | 352404 (29%) | 394819 (31%) | 2422 (6.1%) | 2467 (6.5%) | 2594 (7.1%) | 2625 (7.6%) | 2673 (8.1%) |
| Pressure ulcers, Weight loss | 42157 (4%) | 59180 (5%) | 73054 (6%) | 80767 (7%) | 91152 (7%) | 501 (1.3%) | 610 (1.6%) | 532 (1.4%) | 540 (1.6%) | 557 (1.7%) |
| Incontinence | 25483 (2%) | 29669 (2%) | 32990 (3%) | 39046 (3%) | 49472 (4%) | 211 (0.5%) | 307 (0.8%) | 281 (0.8%) | 265 (0.8%) | 272 (0.8%) |
| Dependence and care | 45667 (4%) | 34745 (3%) | 25496 (2%) | 28474 (2%) | 35131 (3%) | 1218 (3.1%) | 1371 (3.6%) | 1208 (3.3%) | 1112 (3.2%) | 1347 (4.1%) |
| Anxiety and Depression | 88724 (7%) | 116620 (10%) | 141544 (12%) | 162342 (13%) | 195682 (15%) | 1822 (4.6%) | 2346 (6.1%) | 2876 (7.8%) | 3182 (9.2%) | 3690 (11.2%) |

CABG: coronary artery bypass grafting; RCS: Royal College of Surgeons

| **Supplementary Table S3**. Comorbidities (Charlson index RCS) and frailty of admitted heart valve diseases patients and those undergoing heart valve surgery in England 2010-2019 | | | | | | | | | | |
| --- | --- | --- | --- | --- | --- | --- | --- | --- | --- | --- |
|  | Admitted Heart Valve Diseases Patients | | | | | Heart Valve Surgery Patients | | | | |
|  | 2010-2011 | 2012-2013 | 2014-2015 | 2016-2017 | 2018-2019 | 2010-2011 | 2012-2013 | 2014-2015 | 2016-2017 | 2018-2019 |
| N | 251730 | 288038 | 338754 | 394909 | 490981 | 25000 | 26722 | 30067 | 32124 | 35452 |
| Charlson index (RCS) |  |  |  |  |  |  |  |  |  |  |
| 0 | 53623 (21%) | 53586 (19%) | 55079 (16%) | 57181 (14%) | 63878 (13%) | 6104 (24%) | 5691 (21%) | 5849 (19%) | 5279 (16%) | 4665 (13%) |
| 1 | 76939 (31%) | 82843 (29%) | 91256 (27%) | 101406 (26%) | 118549 (24%) | 8324 (33%) | 8781 (33%) | 9417 (31%) | 9424 (29%) | 9669 (27%) |
| 2 | 62905 (25%) | 73054 (25%) | 85469 (25%) | 100000 (25%) | 123706 (25%) | 6060 (24%) | 6731 (25%) | 7640 (25%) | 8572 (27%) | 9709 (27%) |
| 3+ | 58263 (23%) | 78555 (27%) | 106950 (32%) | 136322 (35%) | 184848 (38%) | 4512 (18%) | 5519 (21%) | 7161 (24%) | 8849 (28%) | 11409 (32%) |
| Malignancy | 20722 (8%) | 25149 (9%) | 30924 (9%) | 38590 (10%) | 50825 (10%) | 1005 (4.0%) | 1085 (4.1%) | 1339 (4.5%) | 1630 (5.1%) | 1969 (5.6%) |
| Metastatic solid tumour | 4375 (2%) | 5551 (2%) | 7143 (2%) | 9100 (2%) | 12521 (3%) | 83 (0.3%) | 113 (0.4%) | 142 (0.5%) | 166 (0.5%) | 237 (0.7%) |
| Myocardial infarction | 15523 (6%) | 33747 (12%) | 62786 (19%) | 72567 (18%) | 90752 (18%) | 1692 (7%) | 2789 (10%) | 4531 (15%) | 4836 (15%) | 5243 (15%) |
| Congestive cardiac failure | 103734 (41%) | 123469 (43%) | 150499 (44%) | 185410 (47%) | 239824 (49%) | 10629 (43%) | 11834 (44%) | 13668 (45%) | 16364 (51%) | 20173 (57%) |
| Peripheral vascular disease | 29636 (12%) | 35867 (12%) | 43713 (13%) | 53306 (13%) | 71782 (15%) | 4927 (20%) | 5632 (21%) | 6637 (22%) | 7932 (25%) | 9810 (28%) |
| Cerebrovascular disease | 27152 (11%) | 31634 (11%) | 38665 (11%) | 47854 (12%) | 65230 (13%) | 1866 (7.5%) | 2164 (8.1%) | 2695 (9.0%) | 3249 (10.1%) | 4077 (11.5%) |
| Dementia | 15377 (6%) | 21146 (7%) | 28635 (8%) | 35294 (9%) | 46242 (9%) | 145 (0.6%) | 210 (0.8%) | 270 (0.9%) | 414 (1.3%) | 500 (1.4%) |
| Chronic pulmonary disease | 70101 (28%) | 84400 (29%) | 103538 (31%) | 126863 (32%) | 163286 (33%) | 6616 (26%) | 7436 (28%) | 8628 (29%) | 9534 (30%) | 10819 (31%) |
| Rheumatological disease | 12823 (5%) | 15756 (5%) | 19501 (6%) | 24838 (6%) | 33408 (7%) | 1027 (4.1%) | 1226 (4.6%) | 1525 (5.1%) | 1735 (5.4%) | 1935 (5.5%) |
| Liver disease | 5975 (2%) | 7810 (3%) | 10307 (3%) | 13960 (4%) | 22279 (5%) | 459 (1.8%) | 560 (2.1%) | 682 (2.3%) | 850 (2.6%) | 1391 (3.9%) |
| Diabetes mellitus | 51403 (20%) | 63148 (22%) | 78989 (23%) | 97437 (25%) | 128439 (26%) | 4673 (19%) | 5173 (19%) | 5984 (20%) | 6777 (21%) | 7812 (22%) |
| Hemiplegia or paraplegia | 4168 (2%) | 4509 (2%) | 5711 (2%) | 8007 (2%) | 10883 (2%) | 233 (0.9%) | 253 (0.9%) | 318 (1.1%) | 374 (1.2%) | 462 (1.3%) |
| Renal disease | 48624 (19%) | 60528 (21%) | 76788 (23%) | 96327 (24%) | 129777 (26%) | 2875 (11.5%) | 3127 (11.7%) | 3860 (12.8%) | 4854 (15.1%) | 6519 (18.4%) |
| Frailty syndrome | 100819 (40%) | 128714 (45%) | 164310 (49%) | 203442 (52%) | 270191 (55%) | 5409 (22%) | 6850 (26%) | 8345 (28%) | 10483 (33%) | 12981 (37%) |
| Mental | 28294 (11%) | 39798 (14%) | 54427 (16%) | 69903 (18%) | 93246 (19%) | 1218 (4.9%) | 1714 (6.4%) | 2088 (6.9%) | 2846 (8.9%) | 3535 (10.0%) |
| Mobility problems | 8614 (3%) | 19358 (7%) | 30478 (9%) | 44417 (11%) | 66766 (14%) | 291 (1.2%) | 550 (2.1%) | 754 (2.5%) | 1312 (4.1%) | 1833 (5.2%) |
| Falls and Fractures | 67409 (27%) | 84347 (29%) | 107438 (32%) | 131562 (33%) | 176839 (36%) | 2406 (10%) | 2816 (11%) | 3584 (12%) | 4377 (14%) | 5408 (15%) |
| Pressure ulcers, Weight loss | 11764 (5%) | 18677 (6%) | 26362 (8%) | 32882 (8%) | 44304 (9%) | 579 (2.3%) | 761 (2.8%) | 897 (3.0%) | 1041 (3.2%) | 1281 (3.6%) |
| Incontinence | 6247 (2%) | 8135 (3%) | 10738 (3%) | 14602 (4%) | 22040 (4%) | 201 (0.8%) | 325 (1.2%) | 353 (1.2%) | 453 (1.4%) | 514 (1.4%) |
| Dependence and care | 11366 (5%) | 9282 (3%) | 8375 (2%) | 11248 (3%) | 16523 (3%) | 620 (2.5%) | 695 (2.6%) | 659 (2.2%) | 788 (2.5%) | 1059 (3.0%) |
| Anxiety and Depression | 17947 (7%) | 25723 (9%) | 36304 (11%) | 48459 (12%) | 69717 (14%) | 1158 (4.6%) | 1678 (6.3%) | 2246 (7.5%) | 2992 (9.3%) | 3945 (11.1%) |

RCS: Royal College of Surgeons

**Supplementary Figure S1.** Number and Standardised Rates^*^ of (A) CABG and (B) Heart Valve Surgery in England, UK 2010-2019

| **(A)** | **(B)** |
| --- | --- |
| 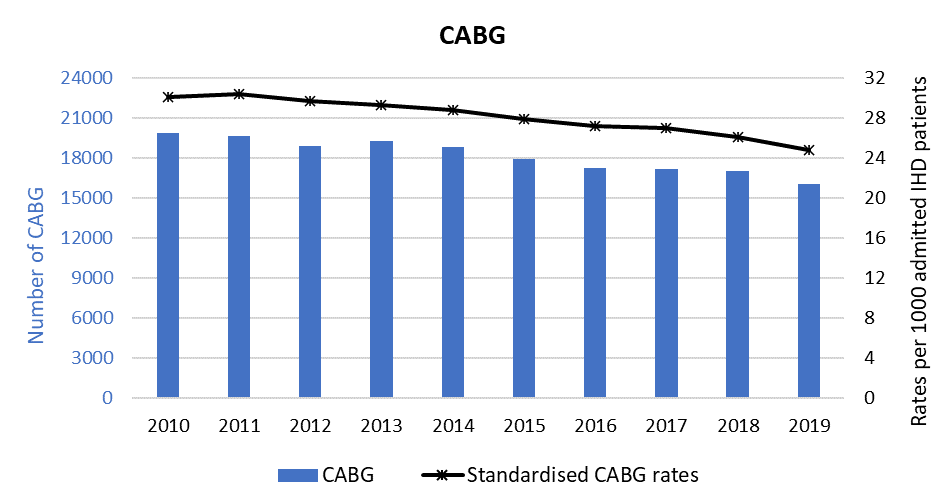 | 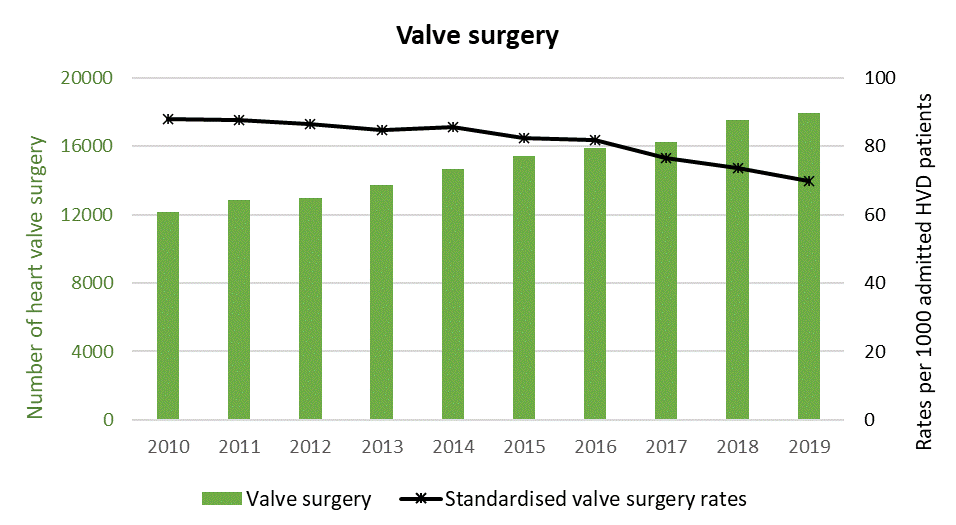 |

**^*^**Standardised rates (by age, Charlson comorbidity index and frailty status) of CABG expressed as per 1000 admitted ischaemic heart disease (IHD) patients and Standardised rates of valve surgery expressed as per 1000 admitted heart valve diseases (HVD) patients

**Supplementary Figure S2.** Standardised rates of CABG per 1000 admitted patients with ischaemic heart disease (A, B, C) and Standardised rates of valve surgery per 1000 admitted patients with heart valves disease (D, E, F) by sex, ethnicity and socio-economic quintile, 2010-2019 (age, comorbidity index, and frailty standardised)

| A | B | C |
| --- | --- | --- |
| 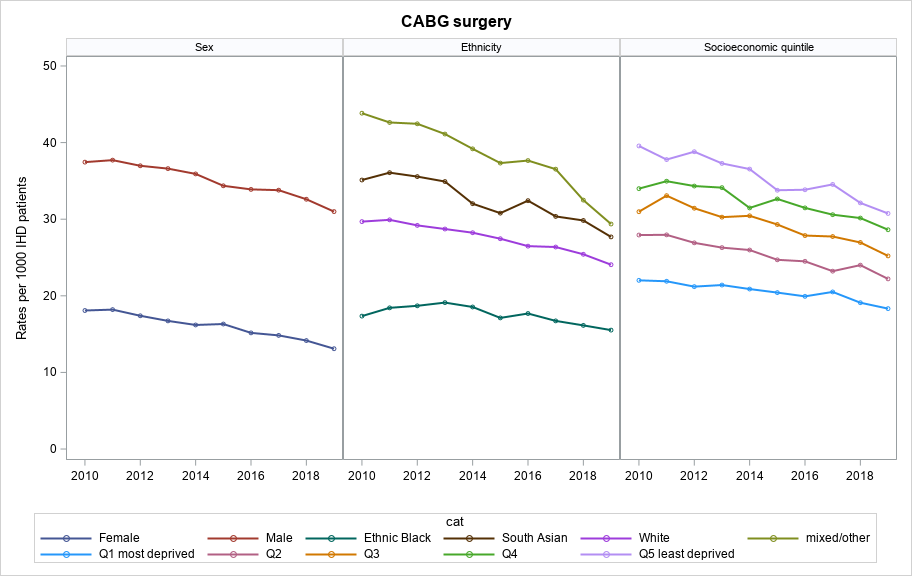 | 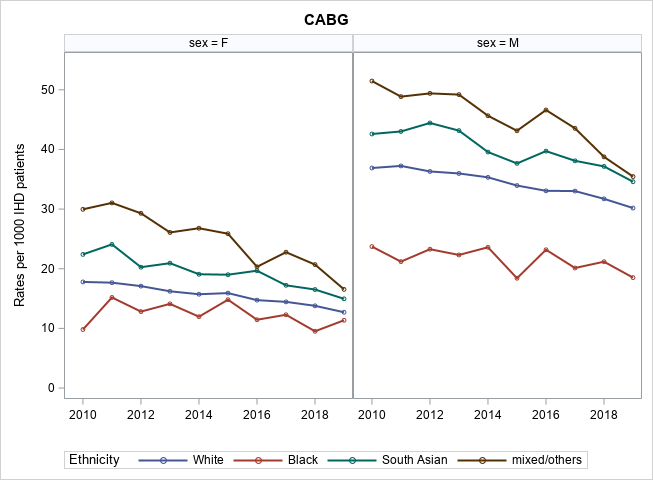 | 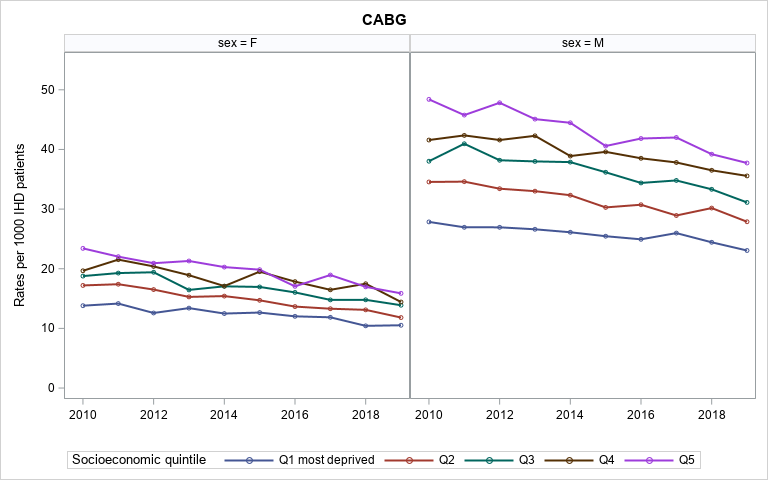 |
| D | E | F |
| 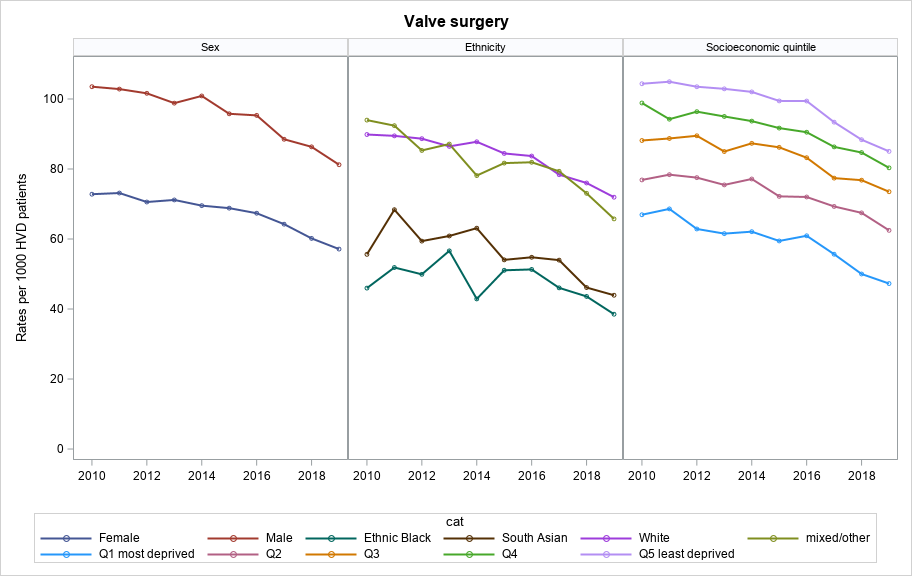 | 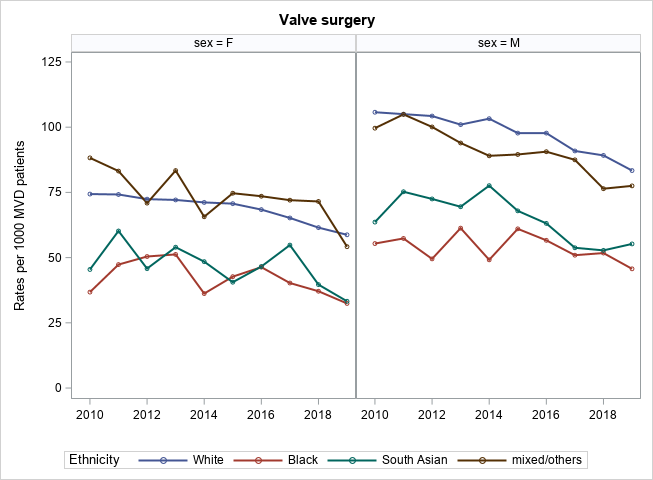 | 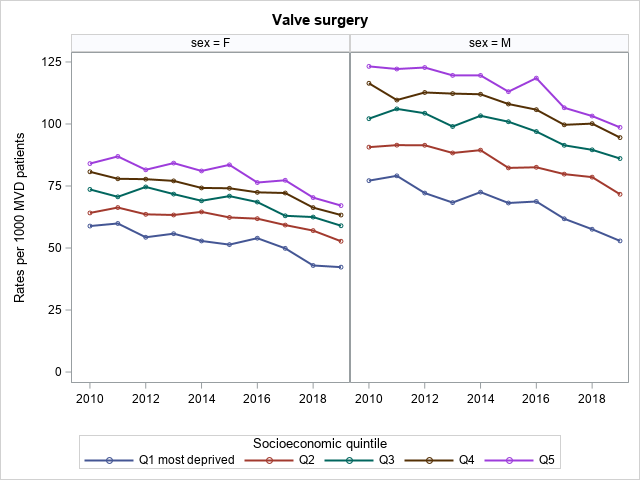 |
| Standardised by age, Charlson comorbidity index and frailty status.  IHD: ischaemic heart disease; HVD: heart valve disease | | |

**Supplementary Table S4.** Characteristics of admitted patients with ischaemic heart disease (IHD) and those undergoing CABG in England, 2010-2019

| **A** | **Admitted Ischaemic Heart Disease (IHD) Patients** | | | | | **CABG Patients** | | | | |
| --- | --- | --- | --- | --- | --- | --- | --- | --- | --- | --- |
|  | 2010-2011 | 2012-2013 | 2014-2015 | 2016-2017 | 2018-2019 | 2010-2011 | 2012-2013 | 2014-2015 | 2016-2017 | 2018-2019 |
| N | 1190870 | 1206648 | 1224683 | 1234160 | 1285859 | 39511 | 38245 | 36774 | 34484 | 33031 |
| per 1000 IHD patients | |  |  |  |  | 33.2 | 31.7 | 30.0 | 27.9 | 25.7 |
| Age mean (SD) | 73.4 (12.7) | 73.6 (12.7) | 73.9 (12.8) | 74.1 (12.7) | 74.1 (12.8) | 67.6 (10.1) | 67.6 (10.1) | 67.6 (10.0) | 67.6 (9.8) | 67.2 (9.7) |
| Age 18-44 | 25793 (2%) | 24720 (2%) | 23934 (2%) | 22719 (2%) | 23850 (2%) | 718 (2%) | 634 (2%) | 608 (2%) | 517 (1%) | 493 (1%) |
| 45-54 | 82746 (7%) | 83475 (7%) | 82549 (7%) | 80521 (7%) | 81908 (6%) | 3831 (10%) | 3725 (10%) | 3567 (10%) | 3171 (9%) | 3073 (9%) |
| 55-64 | 174040 (15%) | 168165 (14%) | 167440 (14%) | 169227 (14%) | 181235 (14%) | 9647 (24%) | 9201 (24%) | 8747 (24%) | 8545 (25%) | 8575 (26%) |
| 65-74 | 287945 (24%) | 290863 (24%) | 291633 (24%) | 295396 (24%) | 306347 (24%) | 14299 (36%) | 13989 (37%) | 13780 (37%) | 13010 (38%) | 12604 (38%) |
| 75+ | 620346 (52%) | 639425 (53%) | 659127 (54%) | 666297 (54%) | 692519 (54%) | 11016 (28%) | 10696 (28%) | 10072 (27%) | 9241 (27%) | 8286 (25%) |
| Sex - male | 693364 (58%) | 711528 (59%) | 728807 (60%) | 741158 (60%) | 779001 (61%) | 31240 (79%) | 30571 (80%) | 29494 (80%) | 27975 (81%) | 27073 (82%) |
| Ethnicity |  |  |  |  |  |  |  |  |  |  |
| White | 1089943 (93%) | 1097440 (92%) | 1104631 (92%) | 1101380 (91%) | 1132799 (90%) | 35257 (90%) | 33739 (89%) | 32229 (89%) | 29564 (88%) | 27716 (87%) |
| Black | 13244 (1%) | 14697 (1%) | 15179 (1%) | 16548 (1%) | 18864 (2%) | 276 (1%) | 329 (1%) | 313 (1%) | 310 (1%) | 321 (1%) |
| South Asian | 47657 (4%) | 51104 (4%) | 54835 (5%) | 58297 (5%) | 63943 (5%) | 2286 (6%) | 2402 (6%) | 2282 (6%) | 2322 (7%) | 2332 (7%) |
| mixed/others | 25620 (2%) | 28120 (2%) | 30906 (3%) | 33708 (3%) | 39630 (3%) | 1435 (4%) | 1500 (4%) | 1509 (4%) | 1558 (5%) | 1551 (5%) |
| Socio-economic quintile | | |  |  |  |  |  |  |  |  |
| Q1 most deprived | 276292 (23%) | 275079 (23%) | 276249 (23%) | 273311 (22%) | 281462 (22%) | 7038 (18%) | 6715 (18%) | 6467 (18%) | 6150 (18%) | 5891 (18%) |
| Q2 | 248257 (21%) | 251025 (21%) | 254179 (21%) | 254109 (21%) | 262999 (21%) | 7649 (20%) | 7234 (19%) | 6910 (19%) | 6335 (19%) | 6267 (19%) |
| Q3 | 241270 (20%) | 245010 (20%) | 248799 (20%) | 252161 (21%) | 262965 (21%) | 8215 (21%) | 7945 (21%) | 7673 (21%) | 7108 (21%) | 6779 (21%) |
| Q4 | 223215 (19%) | 228341 (19%) | 232822 (19%) | 236772 (19%) | 247876 (19%) | 8141 (21%) | 8047 (21%) | 7699 (21%) | 7259 (21%) | 7018 (22%) |
| Q5 least deprived | 192884 (16%) | 198614 (17%) | 203910 (17%) | 209228 (17%) | 221162 (17%) | 7717 (20%) | 7655 (20%) | 7257 (20%) | 6982 (21%) | 6558 (20%) |
| Charlson index (RCS) |  |  |  |  |  |  |  |  |  |  |
| 0 | 320424 (27%) | 259390 (21%) | 193816 (16%) | 177623 (14%) | 169583 (13%) | 10991 (28%) | 9102 (24%) | 7596 (21%) | 6247 (18%) | 5175 (16%) |
| 1 | 392838 (33%) | 372893 (31%) | 342830 (28%) | 326788 (26%) | 320892 (25%) | 13408 (34%) | 12596 (33%) | 11737 (32%) | 10431 (30%) | 9348 (28%) |
| 2 | 266377 (22%) | 294140 (24%) | 316602 (26%) | 317139 (26%) | 324616 (25%) | 8599 (22%) | 9137 (24%) | 9155 (25%) | 8843 (26%) | 8779 (27%) |
| 3+ | 211231 (18%) | 280225 (23%) | 371435 (30%) | 412610 (33%) | 470768 (37%) | 6513 (16%) | 7410 (19%) | 8286 (23%) | 8963 (26%) | 9729 (29%) |
| Frailty syndrome | 427416 (36%) | 480077 (40%) | 532715 (43%) | 576561 (47%) | 642477 (50%) | 6747 (17%) | 7665 (20%) | 8099 (22%) | 8637 (25%) | 9373 (28%) |

Ethnicity missing in n=103,675 (1.7%) and socio-economic quintile missing in n=44,228 (0.7%)

Abbreviations: CABG, coronary artery bypass grafting; IHD: ischaemic heart disease; Q, quintile; SD, standard deviation

**Supplementary Table S5.** Characteristics of admitted patients with heart valve diseases (HVD) and those undergoing valve surgery in England, 2010-2019

| **B** | **Admitted Heart Valve Diseases (HVD) Patients** | | | | | **Valve Surgery Patients** | | | | |
| --- | --- | --- | --- | --- | --- | --- | --- | --- | --- | --- |
|  | 2010-2011 | 2012-2013 | 2014-2015 | 2016-2017 | 2018-2019 | 2010-2011 | 2012-2013 | 2014-2015 | 2016-2017 | 2018-2019 |
| N | 251730 | 288038 | 338754 | 394909 | 490981 | 25000 | 26722 | 30067 | 32124 | 35452 |
| per 1000 HVD patients | |  |  |  |  | 99.3 | 92.8 | 88.8 | 81.3 | 72.2 |
| Age mean (SD) | 75.2 (13.8) | 75.6 (13.8) | 75.9 (13.8) | 76.0 (13.8) | 75.9 (13.8) | 69.1 (12.9) | 69.5 (13.0) | 69.9 (13.0) | 70.7 (13.1) | 70.9 (13.1) |
| Age 18-44 | 9869 (4%) | 10949 (4%) | 12357 (4%) | 13923 (4%) | 16999 (3%) | 1371 (5%) | 1471 (6%) | 1562 (5%) | 1544 (5%) | 1632 (5%) |
| 45-54 | 11558 (5%) | 12930 (4%) | 15459 (5%) | 17903 (5%) | 22740 (5%) | 1732 (7%) | 1751 (7%) | 2050 (7%) | 2025 (6%) | 2324 (7%) |
| 55-64 | 24896 (10%) | 26395 (9%) | 29874 (9%) | 35644 (9%) | 46040 (9%) | 4181 (17%) | 4052 (15%) | 4280 (14%) | 4501 (14%) | 4907 (14%) |
| 65-74 | 50913 (20%) | 57348 (20%) | 66402 (20%) | 77829 (20%) | 96371 (20%) | 7820 (31%) | 8438 (32%) | 9400 (31%) | 9676 (30%) | 10368 (29%) |
| 75+ | 154494 (61%) | 180416 (63%) | 214662 (63%) | 249610 (63%) | 308831 (63%) | 9896 (40%) | 11010 (41%) | 12775 (42%) | 14378 (45%) | 16221 (46%) |
| Sex - male | 121330 (48%) | 140024 (49%) | 165799 (49%) | 196387 (50%) | 247473 (50%) | 15267 (61%) | 16378 (61%) | 18606 (62%) | 19814 (62%) | 22257 (63%) |
| Ethnicity |  |  |  |  |  |  |  |  |  |  |
| White | 233019 (94%) | 265460 (93%) | 310494 (93%) | 357753 (93%) | 438369 (92%) | 23367 (94%) | 24912 (94%) | 27918 (94%) | 29455 (93%) | 32146 (93%) |
| Black | 3862 (2%) | 4767 (2%) | 5580 (2%) | 7315 (2%) | 10051 (2%) | 273 (1%) | 318 (1%) | 342 (1%) | 424 (1%) | 476 (1%) |
| South Asian | 6596 (3%) | 7904 (3%) | 9584 (3%) | 11855 (3%) | 16465 (3%) | 572 (2%) | 629 (2%) | 709 (2%) | 795 (3%) | 863 (2%) |
| mixOth | 5224 (2%) | 6390 (2%) | 7672 (2%) | 9831 (3%) | 13815 (3%) | 654 (3%) | 727 (3%) | 799 (3%) | 936 (3%) | 1072 (3%) |
| Socio-economic quintile | |  |  |  |  |  |  |  |  |  |
| Q1 most deprived | 48202 (19%) | 55475 (19%) | 65245 (19%) | 76122 (19%) | 95079 (20%) | 3893 (16%) | 3919 (15%) | 4388 (15%) | 4752 (15%) | 4907 (14%) |
| Q2 | 49647 (20%) | 56214 (20%) | 65148 (19%) | 75867 (19%) | 93961 (19%) | 4374 (18%) | 4661 (18%) | 5138 (18%) | 5520 (18%) | 6193 (18%) |
| Q3 | 52812 (21%) | 59627 (21%) | 69166 (21%) | 80873 (21%) | 100006 (21%) | 5181 (21%) | 5565 (21%) | 6204 (21%) | 6596 (21%) | 7453 (21%) |
| Q4 | 51161 (21%) | 59150 (21%) | 69773 (21%) | 81495 (21%) | 100801 (21%) | 5474 (22%) | 6018 (23%) | 6705 (23%) | 7198 (23%) | 8129 (23%) |
| Q5 least deprived | 47558 (19%) | 55028 (19%) | 66199 (20%) | 76858 (20%) | 96336 (20%) | 5502 (23%) | 5991 (23%) | 6903 (24%) | 7384 (23%) | 8086 (23%) |
| Charlson index (R |  |  |  |  |  |  |  |  |  |  |
| 0 | 53623 (21%) | 53586 (19%) | 55079 (16%) | 57181 (14%) | 63878 (13%) | 6104 (24%) | 5691 (21%) | 5849 (19%) | 5279 (16%) | 4665 (13%) |
| 1 | 76939 (31%) | 82843 (29%) | 91256 (27%) | 101406 (26%) | 118549 (24%) | 8324 (33%) | 8781 (33%) | 9417 (31%) | 9424 (29%) | 9669 (27%) |
| 2 | 62905 (25%) | 73054 (25%) | 85469 (25%) | 100000 (25%) | 123706 (25%) | 6060 (24%) | 6731 (25%) | 7640 (25%) | 8572 (27%) | 9709 (27%) |
| 3+ | 58263 (23%) | 78555 (27%) | 106950 (32%) | 136322 (35%) | 184848 (38%) | 4512 (18%) | 5519 (21%) | 7161 (24%) | 8849 (28%) | 11409 (32%) |
| Frailty syndrome | 100819 (40%) | 128714 (45%) | 164310 (49%) | 203442 (52%) | 270191 (55%) | 5409 (22%) | 6850 (26%) | 8345 (28%) | 10483 (33%) | 12981 (37%) |

Ethnicity missing in n=32,406 (1.8%) and socio-economic quintile missing in n=16,609 (0.9%)

Abbreviations: HVD, heart valve disease; Q, quintile; SD, standard deviation

**Supplementary Table S6.** Outcomes of patients undergoing CABG and valve surgery in 2010-2019 by sex, ethnic group and socio-economic quintile

|  | 2010-2014 | | | | | 2015-2019 | | | | |
| --- | --- | --- | --- | --- | --- | --- | --- | --- | --- | --- |
|  | CABG alone | Valve surgery alone | Aortic valve | Mitral valve | Concomitant CABG & valve surgery | CABG alone | Valve surgery alone | Aortic valve | Mitral valve | Concomitant CABG & valve surgery |
| No of patients | 76604 | 46308 | 30397 | 9331 | 20192 | 67416 | 63491 | 45094 | 10785 | 18129 |
| **In-hospital mortality** |  |  |  |  |  |  |  |  |  |  |
| Sex F | 2.7% | 3.4% | 2.9% | 3.2% | 6.9% | 2.2% | 2.6% | 2.0% | 2.5% | 6.4% |
| M | 1.5% | 2.6% | 2.2% | 2.1% | 4.8% | 1.2% | 2.1% | 1.7% | 1.6% | 3.9% |
| Ethnicity |  |  |  |  |  |  |  |  |  |  |
| White | 1.6% | 2.9% | 2.4% | 2.6% | 5.2% | 1.3% | 2.2% | 1.8% | 1.9% | 4.3% |
| Black | 2.9% | 4.8% | 4.7% | 3.7% | 4.6% | 3.1% | 3.1% | 2.6% | 2.7% | 5.5% |
| South Asian | 2.2% | 4.7% | 4.2% | 3.3% | 8.8% | 1.7% | 3.4% | 2.3% | 2.4% | 8.9% |
| mixed/ others | 1.7% | 2.7% | 2.4% | 1.8% | 7.2% | 1.4% | 2.2% | 2.0% | 1.5% | 5.6% |
| Socio-economic quintile |  |  |  |  |  |  |  |  |  |  |
| Q1 most deprived | 1.8% | 3.2% | 2.8% | 3.0% | 6.6% | 1.7% | 2.6% | 2.0% | 2.4% | 5.4% |
| Q2 | 1.8% | 3.2% | 2.5% | 3.1% | 5.2% | 1.3% | 2.6% | 2.3% | 1.6% | 4.7% |
| Q3 | 1.8% | 2.8% | 2.4% | 2.3% | 5.8% | 1.2% | 2.3% | 1.7% | 2.1% | 4.4% |
| Q4 | 1.6% | 2.8% | 2.5% | 2.5% | 4.5% | 1.5% | 1.9% | 1.5% | 1.6% | 4.3% |
| Q5 least deprived | 1.5% | 3.0% | 2.4% | 2.4% | 5.4% | 1.3% | 2.3% | 1.8% | 2.2% | 3.8% |
| **1-year Mortality (all-cause)** | |  |  |  |  |  |  |  |  |  |
| Sex F | 5.4% | 8.0% | 7.7% | 6.5% | 11.9% | 4.6% | 7.4% | 7.4% | 5.3% | 11.4% |
| M | 3.5% | 7.3% | 7.4% | 4.6% | 10.0% | 3.2% | 7.6% | 8.1% | 4.6% | 8.4% |
| Ethnicity |  |  |  |  |  |  |  |  |  |  |
| White | 3.8% | 7.6% | 7.5% | 5.5% | 10.3% | 3.4% | 7.6% | 7.9% | 4.9% | 9.0% |
| Black | 4.8% | 10.0% | 11.5% | 6.1% | 15.7% | 4.9% | 7.5% | 8.0% | 8.1% | 8.6% |
| South Asian | 4.0% | 8.2% | 8.8% | 5.6% | 12.8% | 3.4% | 6.9% | 6.7% | 5.3% | 12.7% |
| mixed/ others | 3.6% | 5.9% | 5.8% | 4.3% | 10.9% | 2.8% | 5.9% | 7.3% | 3.3% | 8.5% |
| Socio-economic quintile |  |  |  |  |  |  |  |  |  |  |
| Q1 most deprived | 4.1% | 8.0% | 8.0% | 6.8% | 12.1% | 3.9% | 8.3% | 8.2% | 6.0% | 10.6% |
| Q2 | 3.9% | 8.3% | 8.0% | 6.4% | 10.6% | 3.3% | 8.4% | 8.7% | 6.2% | 9.8% |
| Q3 | 4.1% | 7.5% | 7.5% | 5.3% | 10.7% | 3.2% | 7.5% | 7.8% | 4.7% | 9.3% |
| Q4 | 3.6% | 7.4% | 7.5% | 5.1% | 9.6% | 3.3% | 6.9% | 7.4% | 3.8% | 8.5% |
| Q5 least deprived | 3.4% | 7.1% | 7.0% | 4.5% | 10.0% | 3.4% | 7.2% | 7.4% | 4.6% | 7.8% |
| **Cardiovascular-cause hospital readmission at 1 year** | | | |  |  |  |  |  |  |  |
| Sex F | 20.8% | 24.6% | 22.8% | 26.5% | 27.0% | 17.7% | 22.2% | 20.9% | 24.2% | 24.6% |
| M | 16.5% | 25.9% | 24.2% | 27.4% | 26.5% | 15.1% | 24.2% | 22.9% | 25.8% | 23.8% |
| Ethnicity |  |  |  |  |  |  |  |  |  |  |
| White | 17.5% | 25.5% | 23.8% | 27.3% | 26.8% | 15.8% | 23.7% | 22.3% | 25.6% | 24.3% |
| Black | 20.2% | 29.1% | 26.6% | 29.3% | 44.4% | 18.0% | 26.6% | 26.3% | 29.4% | 26.6% |
| South Asian | 15.9% | 22.1% | 19.1% | 25.2% | 23.3% | 14.9% | 21.8% | 21.8% | 19.5% | 22.4% |
| mixed/ others | 16.6% | 23.7% | 20.5% | 24.0% | 25.6% | 15.8% | 22.8% | 20.5% | 24.1% | 20.7% |
| Socio-economic quintile |  |  |  |  |  |  |  |  |  |  |
| Q1 most deprived | 18.0% | 25.5% | 23.4% | 27.8% | 26.7% | 16.2% | 23.1% | 21.5% | 24.8% | 22.8% |
| Q2 | 17.9% | 25.2% | 23.7% | 25.9% | 26.7% | 15.8% | 24.0% | 22.6% | 26.7% | 24.4% |
| Q3 | 18.1% | 26.4% | 24.5% | 28.4% | 28.1% | 15.8% | 24.2% | 22.8% | 26.2% | 25.5% |
| Q4 | 17.1% | 25.2% | 23.5% | 26.9% | 28.2% | 15.4% | 23.6% | 22.1% | 25.3% | 24.2% |
| Q5 least deprived | 16.5% | 26.4% | 24.7% | 27.5% | 25.5% | 15.4% | 23.6% | 22.4% | 24.8% | 24.3% |
| **Hospital admission for heart failure at 1 year** | | |  |  |  |  |  |  |  |  |
| Sex F | 8.0% | 12.0% | 11.0% | 11.4% | 13.8% | 9.5% | 15.0% | 14.4% | 14.7% | 15.6% |
| M | 6.0% | 11.5% | 11.4% | 9.6% | 12.7% | 7.7% | 14.9% | 15.0% | 12.8% | 14.6% |
| Ethnicity |  |  |  |  |  |  |  |  |  |  |
| White | 6.3% | 11.8% | 11.3% | 10.5% | 12.8% | 8.0% | 15.0% | 14.8% | 13.5% | 14.7% |
| Black | 7.7% | 15.0% | 12.6% | 17.1% | 24.1% | 12.5% | 20.3% | 19.0% | 24.4% | 14.8% |
| South Asian | 6.8% | 11.8% | 9.8% | 8.6% | 17.8% | 8.8% | 15.0% | 15.6% | 11.2% | 19.1% |
| mixed/ others | 7.5% | 10.6% | 9.5% | 8.4% | 15.8% | 8.5% | 15.7% | 15.0% | 14.5% | 17.3% |
| Socio-economic quintile |  |  |  |  |  |  |  |  |  |  |
| Q1 most deprived | 7.3% | 14.1% | 13.2% | 14.5% | 15.5% | 9.6% | 17.3% | 17.0% | 16.4% | 16.2% |
| Q2 | 7.0% | 12.6% | 11.9% | 11.2% | 13.8% | 8.3% | 15.7% | 15.1% | 15.3% | 16.2% |
| Q3 | 6.7% | 11.6% | 11.0% | 9.8% | 13.6% | 8.1% | 15.2% | 15.0% | 13.7% | 14.7% |
| Q4 | 5.9% | 11.4% | 11.4% | 9.1% | 13.3% | 7.5% | 14.5% | 14.4% | 12.6% | 14.8% |
| Q5 least deprived | 5.4% | 10.8% | 10.3% | 9.6% | 10.9% | 7.1% | 14.2% | 14.1% | 12.5% | 13.9% |

| **Hospital admission for stroke/TIA at 1 year** | | |  |  |  |  |  |  |  |  |
| --- | --- | --- | --- | --- | --- | --- | --- | --- | --- | --- |
| Sex F | 2.9% | 3.4% | 3.4% | 3.3% | 4.1% | 2.3% | 3.9% | 4.1% | 3.1% | 4.6% |
| M | 1.9% | 3.4% | 3.6% | 2.9% | 4.0% | 1.9% | 3.8% | 4.0% | 2.8% | 4.1% |
| Ethnicity |  |  |  |  |  |  |  |  |  |  |
| White | 2.2% | 3.4% | 3.5% | 3.0% | 4.1% | 2.0% | 3.9% | 4.1% | 2.9% | 4.3% |
| Black | 2.4% | 4.4% | 4.0% | 5.5% | 5.6% | 2.7% | 4.0% | 3.8% | 4.1% | 7.8% |
| South Asian | 1.8% | 3.1% | 3.6% | 2.3% | 4.2% | 2.0% | 3.8% | 4.6% | 2.7% | 3.7% |
| mixed/ others | 2.2% | 3.3% | 2.6% | 4.1% | 3.5% | 2.1% | 3.6% | 3.8% | 3.3% | 3.6% |
| Socio-economic quintile |  |  |  |  |  |  |  |  |  |  |
| Q1 most deprived | 2.2% | 3.4% | 3.1% | 4.1% | 4.6% | 2.1% | 3.6% | 3.9% | 2.9% | 4.1% |
| Q2 | 2.2% | 3.5% | 3.6% | 2.6% | 3.8% | 2.2% | 3.9% | 4.1% | 3.2% | 4.9% |
| Q3 | 2.2% | 3.5% | 3.5% | 3.4% | 3.9% | 1.9% | 3.9% | 4.1% | 3.2% | 5.0% |
| Q4 | 2.1% | 3.4% | 3.6% | 2.9% | 4.3% | 2.1% | 4.0% | 4.4% | 2.7% | 3.7% |
| Q5 least deprived | 2.0% | 3.5% | 3.8% | 2.6% | 3.9% | 1.8% | 3.9% | 4.1% | 2.7% | 3.8% |

Abbreviations: CABG, Coronary artery bypass grafting; Q, Quintile, TIA, Transient ischaemic attack
